# Limited recovery from post-acute sequelae of SARS-CoV-2 (PASC) at eight months in a prospective cohort

**DOI:** 10.1101/2021.03.29.21254211

**Authors:** DR Darley, GJ Dore, A Byrne, M Plit, BJ Brew, A Kelleher, GV Matthews

**Affiliations:** Department of Thoracic Medicine, St Vincent’s Hospital Darlinghurst, Sydney, Australia; Department of Infectious Diseases, St Vincent’s Hospital Darlinghurst, Sydney, Australia; UNSW Medicine, St Vincent’s Clinical School, University of New South Wales, Sydney, Australia; Department of Neurology and Peter Duncan Neurosciences Unit St Vincent’s Centre for Applied Medical Research, Sydney Australia; Kirby Institute, University of New South Wales, Sydney, Australia; Department of Immunology, St Vincent’s Hospital Darlinghurst, Sydney, Australia

## Abstract

There is increasing recognition of the prolonged illness following acute coronavirus disease 2019 (COVID-1). In a longitudinal cohort of 99 patients, 32% reported persistent symptoms and 19% had Long COVID (Defined as fatigue or dyspnoea or chest tightness) at median 240 days after initial infection. There was no significant improvement in symptoms or measures of health-related quality of life between 4 and 8-month assessments. In multivariable analysis, female gender (OR 3.2, 95%CI 1.3-7.8, p=0.01) and acute COVID-19 hospitalisation (OR 3.8, 95% 1.1-13.6, p=0.04) were independently associated with Long COVID at 8-months. Only 80% patients reported full recovery at 8 months. Further research is required to understand the immunologic correlates of abnormal recovery and the long-term significance.

## Introduction

Global attention is gradually turning to focus on the problem of prolonged illness following acute coronavirus disease 2019 (COVID-19), commonly termed ‘Long COVID’ or Post-Acute Sequelae of SARS-CoV-2 infection (PASC). Whilst an increasing number of reports now recognise this condition, accurate characterisation of its prevalence, clinical features and natural history is complicated by choice of denominator population, lack of case definition and marked self-selection bias. Nevertheless, a picture is emerging of a syndrome characterised predominantly by fatigue, dyspnoea, chest tightness, and ‘brain fog’ present in around 10-30% of individuals at 2-3 months post-acute infection and affecting both those with initial severe illness and those in whom acute infection was mild (1-3).

In April 2020 we commenced a prospective observational cohort study (ADAPT) following all patients with a positive SARS-CoV-2 RNA test through our hospital testing centres. At a median of 79 days post infection, 40% reported persistent symptoms including a high proportion of those managed in the community for initial infection (4). We now report further 8-month post-infection follow-up to characterise persistence of symptoms, measures of health-related quality of life (HRQOL) and recovery, and assess within-individual changes in measures of psychologic and somatic dysfunction.

## Methods

The ADAPT study is a prospective cohort at St Vincent’s Hospital Sydney, with nasopharyngeal swab-confirmed SARS-CoV-2 infection, being followed for up to 24 months (4). Data from the 4- and 8- month assessments for patients with initial positive PCR from 09-Mar-2020 until 28-Apr-2020 were analysed. This cohort includes both patients who were diagnosed at St Vincent’s Hospital testing clinics (internal) and self-referred patients (external). Standardised case report forms were used to collect specific symptoms at 4 and 8-months after infection. Patients were classified as ‘Long COVID’ if ≥ 1 of the following persistent symptoms were reported; fatigue, dyspnoea, or chest pain. The Somatic and Psychologic Health Report-34 item SPHERE-34 is a self-report questionnaire containing 34 questions surveying mental distress and persistent fatigue, validated as a screening tool for mental health in a variety of Australian populations. (5, 6) **(Supplementary File)**. Three sub-scales measuring anxiety-depression, somatic distress, and persistent fatigue are scored to produce total and somatic and psychological subscale scores. Key somatic symptoms such as muscle pain or tiredness after activity, needing to sleep longer or poor sleep, and prolonged tiredness after activity. A visual analogue scale for fatigue (VAS-F) and the Medical Research Council (MRC) dyspnoea scale, to compare breathlessness pre-COVID and at 8-months, were performed (7, 8). Functional recovery was interrogated on a 5-point Likert scale. Data distribution was tested using the Shapiro-Wilk test. Descriptive statistics were summarised by mean and standard deviation (SD) or median with interquartile range (IQR) for continuous variables, and counts (%) for categorical variables. The Wilcoxon matched-pairs signed rank test and Fisher’s exact test were used to compare medians and proportions. Multivariable logistic regression analysis was performed to identify independent predictors of major symptoms at 8-months follow-up. Statistical analyses were performed using Prism Version 8.4.3. Statistical significance was set at a 2-sided level of 0.05.

## Results

A total of 99 patients underwent 8-month assessment at a median 240 (IQR 227-256) days after positive SARS-CoV-2 PCR (**Supplementary Figure 1**). Of these, 66 patients were diagnosed at St Vincent’s Hospital testing clinics and 33 patients were self-referred following external diagnosis. Fifteen patients originally enrolled in ADAPT did not attend (n=6), were lost to follow-up (n=7), or withdrew consent (n=2). The majority (87%) of patients were managed in the community. Median age was 47 (IQR 35-58) with 61% males.

### Prevalence of symptoms at 8-month follow-up

To minimise self-selection bias, prevalence of symptoms at 8 months is reported for patients diagnosed at St Vincent’s Hospital testing clinics only. Twenty-six (40%) patients had one or more symptoms (new or persistent) at 8-months **(Figure 1)**. Most common symptoms were fatigue (23%) and shortness of breath (11%). In a sensitivity analysis, assuming all patients who were lost to follow-up had fully recovered, the proportion of symptomatic patients at month 8 was 32% (26/81) with 19% (15/81) meeting our definition for Long COVID. In all subsequent analyses all patients (St Vincent’s Hospital diagnosed and external) in our cohort were included.

**Figure 1.**
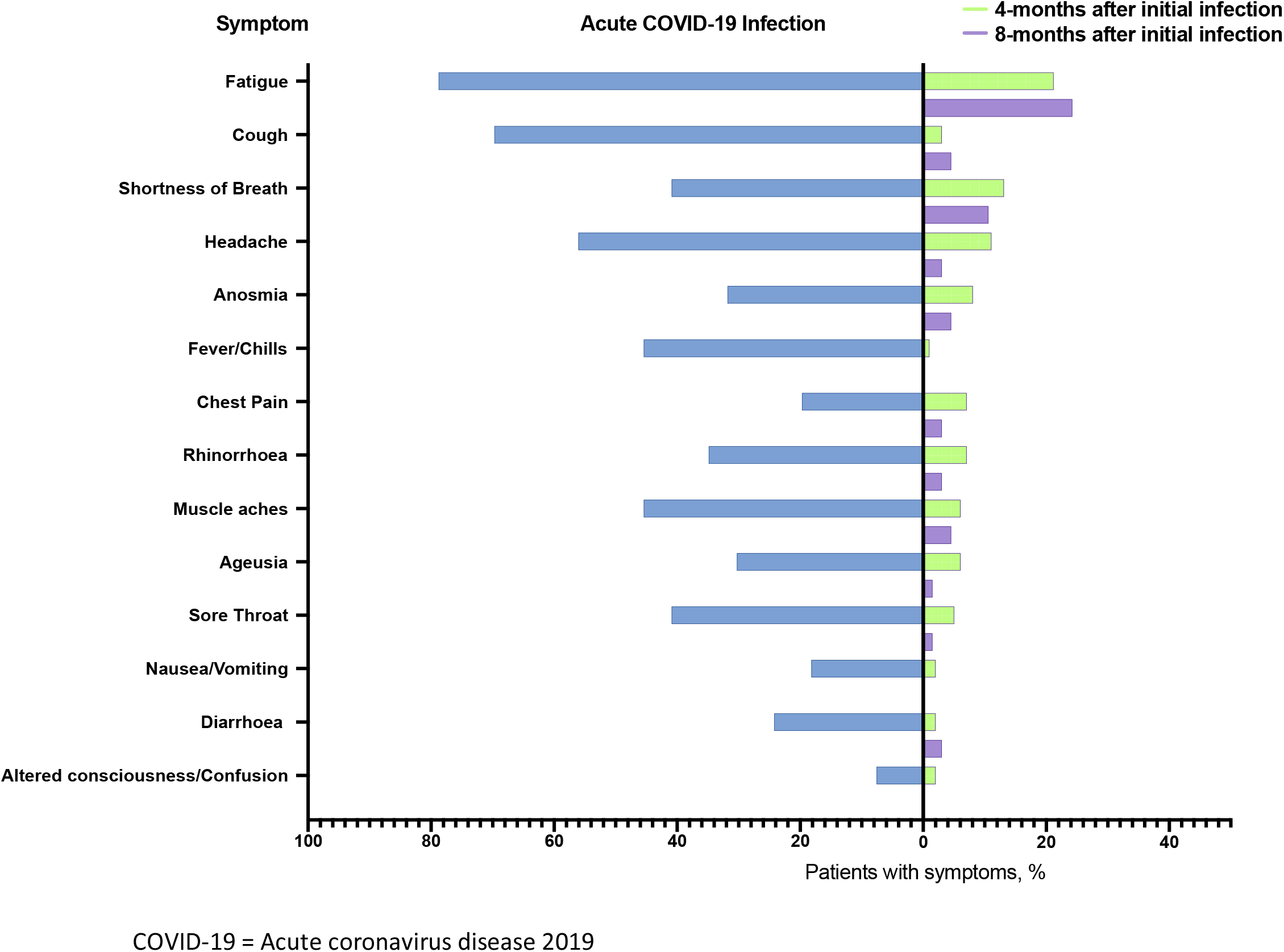
**Proportion of patients with symptoms at acute coronavirus disease 2019 (COVID-19) infection compared with 8 months after assessment (n=66).**

### Presence and predictors of Long COVID symptoms at 8-months follow-up

Ninety-two participants completed a study assessment at both 4- and 8-months. There was no significant difference in the proportion of patients reporting symptoms at 4-months (41/92, 45%) and 8-months (47/92, 51%) (p=0.47). There was no significant difference in the proportion of patients with Long COVID at 4-months (27/92, 29%) compared to 8 months (32/92, 35%) (p=0.53) (**Figure 2**).

**Figure 2.**
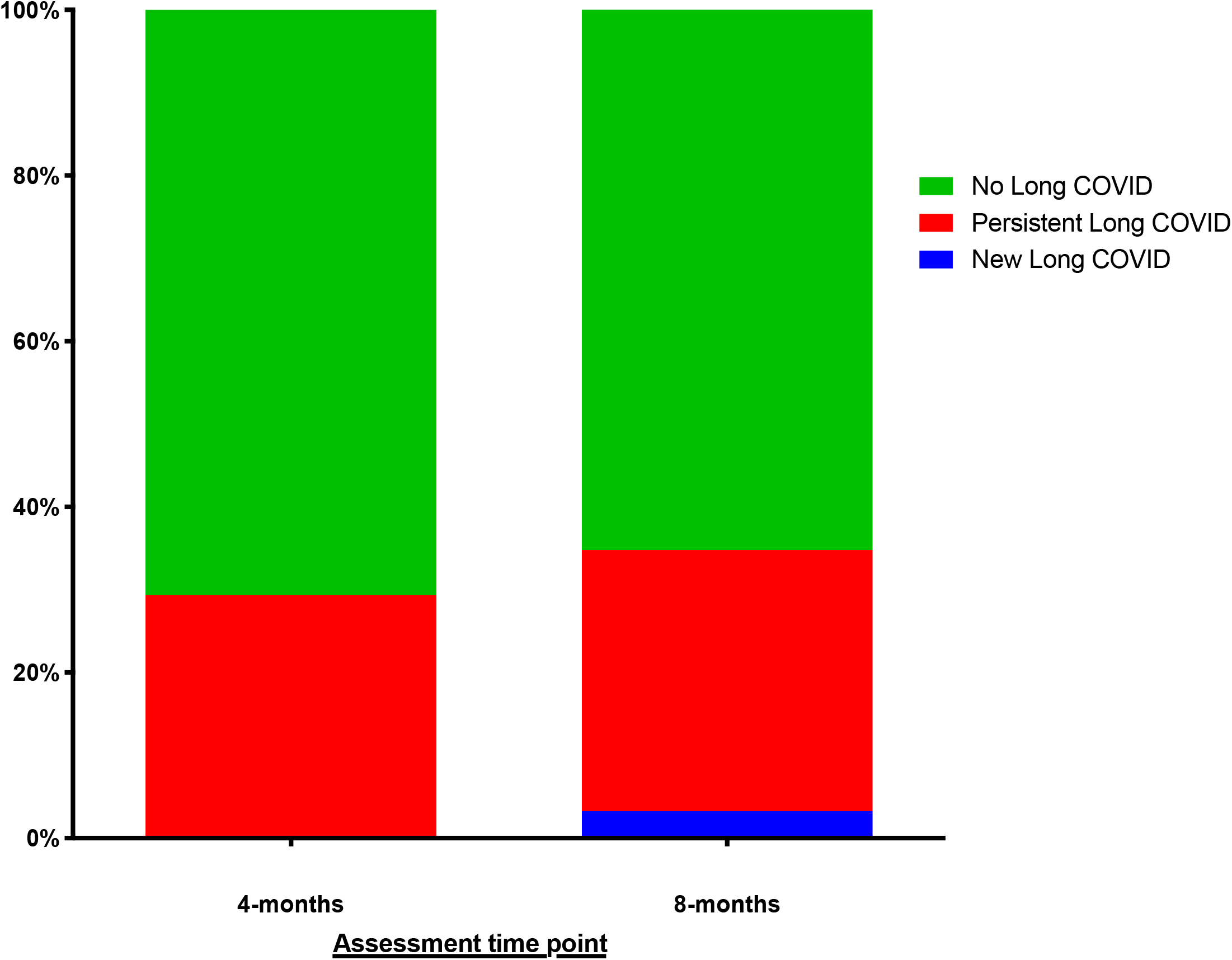
**Stacked bar charts comparing the proportion of patients with Long COVID at 4- and 8-month assessments in n=92 patients. Long COVID definition includes persistent fatigue or shortness of breath or chest tightness/pain.**

In univariable logistic regression analysis, female gender (OR 2.7, 95%CI 1.2-6.3, p=0.02) was associated with major symptoms (Long COVID) at 8-month follow-up. In multivariable analysis, female gender (OR 3.2, 95%CI 1.3-7.8, p=0.01) and acute COVID-19 hospitalisation (OR 3.8, 95% 1.1- 13.6, p=0.04) were both independently associated with Long COVID at 8-month follow-up. The results are summarised in **Table 1**.

**Table 1.** **Results of the univariable and multivariable logistic regression model measuring the association between predictors of interest and risk of Long COVID at 8 months. Variables with univariable p<0.1 were included in the final model. Hospital length of stay was not added to the final model to avoid over-adjustment. Female gender and hospitalisation for initial infection were both independently associated with risk of Long COVID at 8 months.**

| Variable | Univariable Odds Ratio (95%CI) | P value | Multivariable Odds Ratio (95% CI) | P value |
| --- | --- | --- | --- | --- |
| Gender<br>- Female<br>- Male | 2.7 (1.2-6.3)<br>Reference | 0.02 | 3.2 (1.3-7.8)<br>Reference | **0.01** |
| Hospitalisation<br>- Yes<br>- No | 2.9 (0.9-9.4)<br>Reference | 0.09 | 3.8 (1.1-13.6)<br>Reference | **0.04** |
| Age at diagnosis (1-unit) | 1.0 (1.0-1.0) | 0.35 |  |  |
| Hospital Length of Stay | 1.7 (0.9-3.1) | 0.08 |  |  |
| Co-morbidities<br>- Yes<br>- No | 0.9 (0.4-2.2)<br>Reference | 0.87 |  |  |

### HRQOL measures

A total of 97 patients underwent SPHERE-34 testing at 8-month assessment involving measures of both somatic and psychological (cognitive) symptoms: 27% patients reported poor memory a good part of, or most of the time; 33% patients reported poor concentration a good part of, or most of the time; and 18% patients reported feeling lost for the word a good part of, or most of the time.

To understand the evolving nature of these symptoms over time, we further evaluated changes in measurements between 4 and 8 months.

### Longitudinal change in symptoms and mental health between 4- and 8-months follow-up

Complete paired SPHERE-34 at both timepoints, were available in 88 patients (**Table 2**). For each patient, the within-individual change in SPHERE-34 was calculated as the 8-month value minus the 4- month value. The median intersession change in SPHERE-34 total score was 0.0 (IQR -3.8-2.0). There were no significant differences in the median total SPHERE-34 scores between 4-months (4.5, IQR 1.0-12.0) and 8-months (4.0, IQR 0.0-10.0) (p=0.19). There were no significant differences in the mean SOMA scores between 4- and 8-months, nor in the proportion of patients with abnormal SOMA scores. There was a significant difference in the mean PSYCH score between 4-months (1.7, SD 2.4) and 8-months (1.0, SD 1.9) (p=0.03). There was a trend to decreasing proportions of abnormal PSYCH between 4- and 8-months. The results are summarised in **Figure 3A, 3B, 3C**.

**Table 2.** **Comparison of total SPHERE-34 and subscales (SOMA, PSYCH) between the 4- and 8-month assessments in n=88 patients.**

| **<u>Variable</u>** | **4-months** | **8 months** | **P value** |
| --- | --- | --- | --- |
| Median Total SPHERE-34 Score (IQR) | 4.5 (1.0-12.0) | 4.0 (0.0-10.0) | 0.19 |
| Median SOMA Sub-scale Score (IQR) | 1.0 (0.0-4.0) | 1.0 (0.0-4.0) | 0.75 |
| - %-abnormal SOMA (n, %-total) | 32/88 (36.4%) | 34/88 (38.6%) | 0.88 |
| Median PSYCH Sub-scale Score (IQR) | 0.0 (0.0-2.0) | 0.0 (0.0-1.0) | 0.02 |
| - %-abnormal PSYCH (n, %-total) | 30/88 (34.1%) | 21/88 (23.9%) | 0.18 |

**SPHERE-34 = Somatic and Psychologic Health Report-34 item; SOMA = Somatic sub-scale; PSYCH = Psychologic sub-scale**

**Figure 3.**
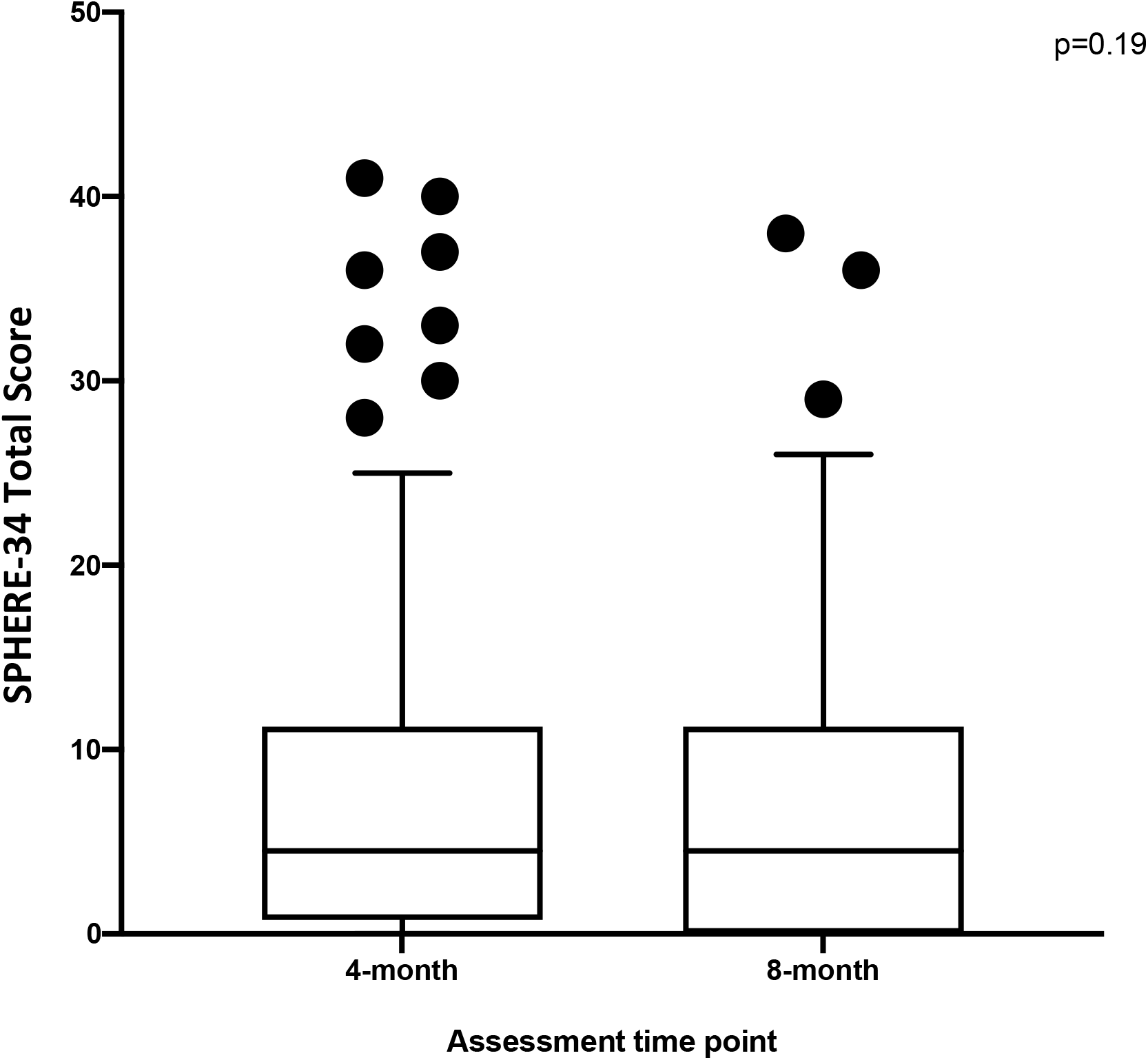
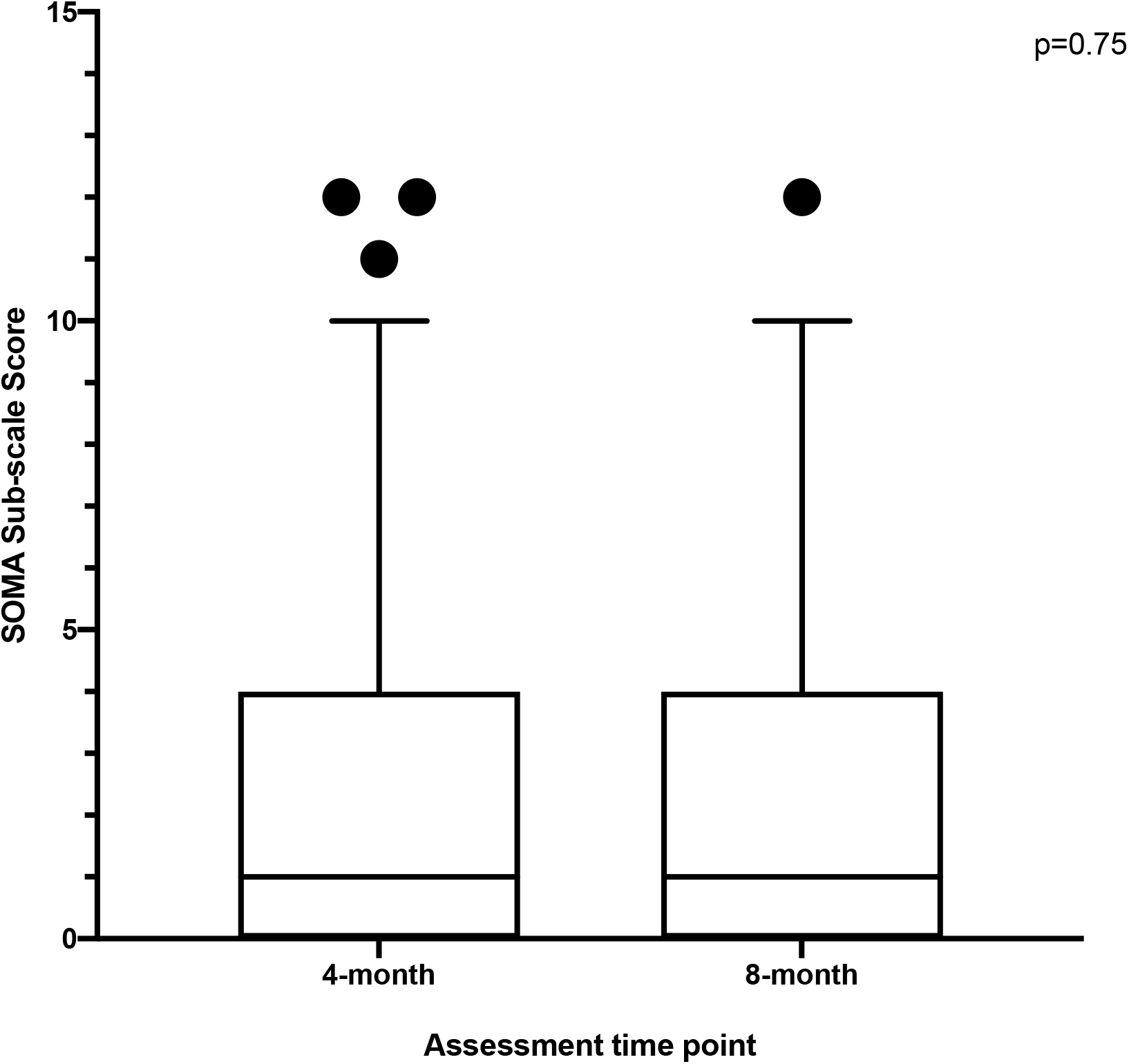
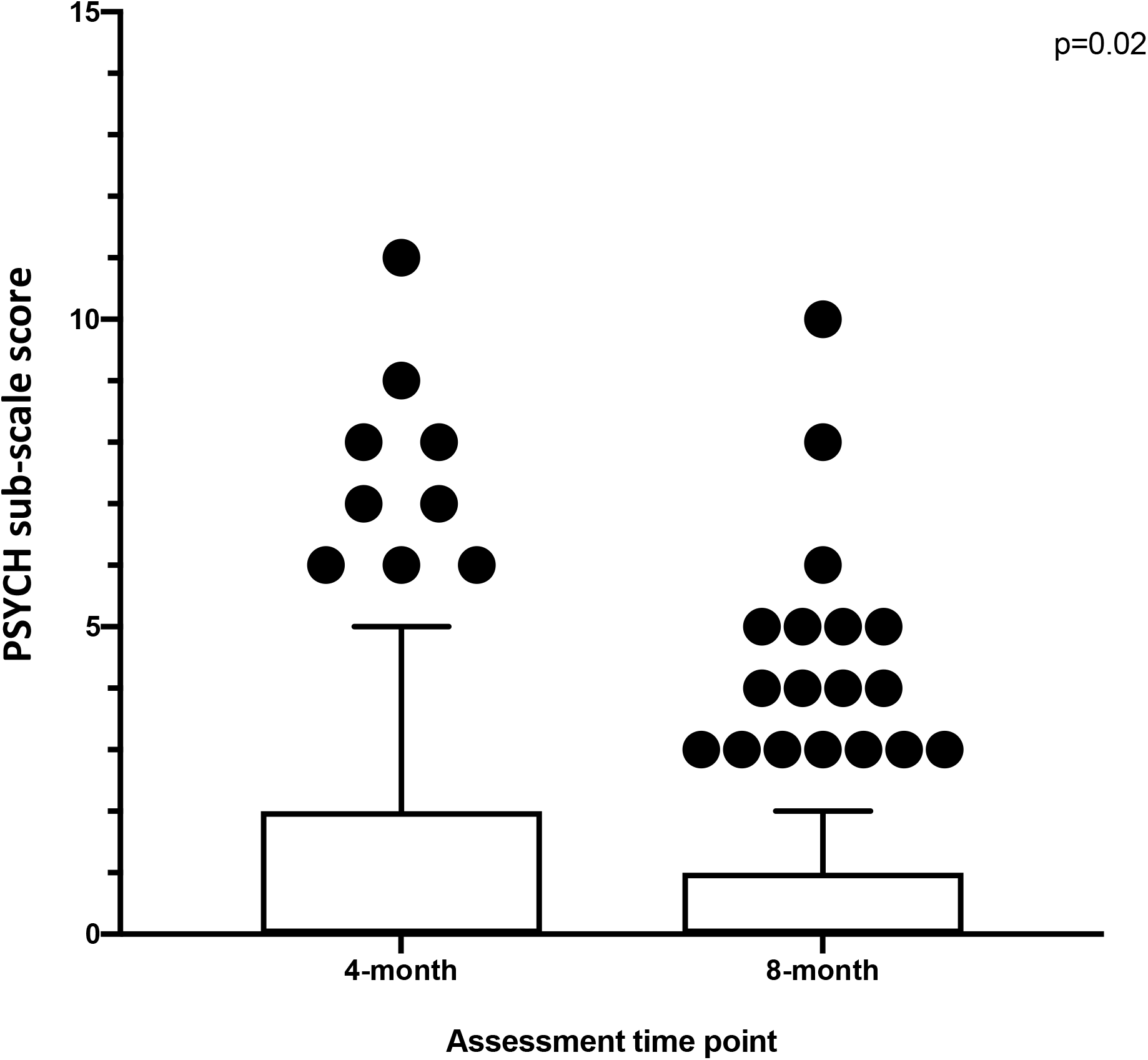
**Tukey box plots for SPHERE-34 at 4- and 8-month assessments in n=88 patients. Figure 3A: Total scores, Figure 3B: Somatic (SOMA) sub-scale, and Figure 3C: Psychologic (PSYCH) sub-scale. Outlier values are represented as black circles.** **Figure 3A. Tukey box plots for SPHERE-34 total scores at 4-month and 8-month assessments** **Figure 3B. Tukey box plots for SOMA sub-scale scores at 4-month and 8-month assessments** **Figure 3C. Tukey box plots for PSYCH sub-scale scores at 4-month and 8-month assessments**

### Functional Outcomes at 8-month assessment

Ninety-eight patients underwent assessment for functional recovery at 8 months. 28/98 (29%) patients reported an increase in dyspnoea, as measured by the MRC dyspnoea scale from pre-COVID levels. The median fatigue visual analogue scale score was 2.0 (IQR 0.38-5.0). In regard to COVID-19 recovery 78/98 (80%) agreed they had fully recovered, 88/98 (90%) agreed they felt confident returning to pre-COVID work, 89/98 (91%) agreed they had returned to usual activities of daily living, while 76/98 (78%) agreed they had returned to normal exercise level (Table 3). In all measures, recovery was significantly lower among patients with Long COVID, with only 54% agreeing that they had recovered from COVID-19 (**Table 3**).

**Table 3.**
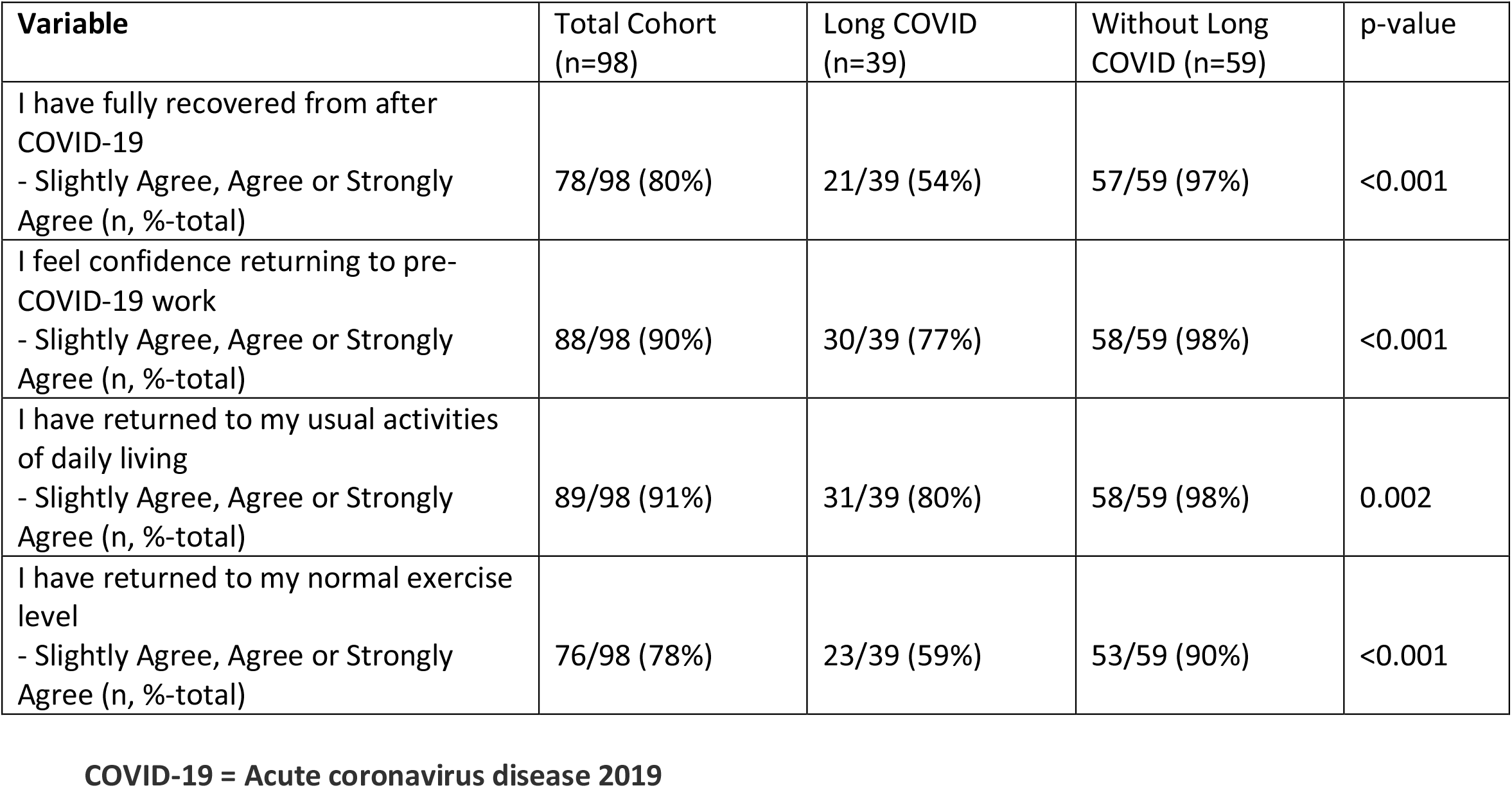
**Comparison of recovery Likert questionnaire in patients with Long COVID at 8-month assessment compared to those without (n=98).**

## Discussion

The spectrum of long-term recovery following SARS-CoV-2 infection remains uncertain. Our study documents the longitudinal nature and prevalence of persistent symptoms and the effect on HRQOL, and delivers potentially concerning findings. At a median of 8 months after infection, even when self-referred patients were excluded, a third had persistent symptoms. A fifth of patients could be classified as having ‘Long COVID’ and there appears to be minimal improvement between 4- and 8- months after infection, including no significant differences in the total scores or somatic sub-scales of SPHERE-34. In the total cohort, we observed a significant difference in SPHERE-34 mean psychologic sub-scales scores indicating some improvement in psychologic symptoms. Concerningly, a considerable proportion of (around 20%) the total cohort did not feel confident returning to pre-COVID work, had not returned to usual activities of daily living or had not returned to normal exercise level.

The aetiology of persisting symptoms following SARS-COV2 infection is likely to be multifactorial and encompassing both prolonged recovery from persisting cardiothoracic damage as has been demonstrated by us and others (4, 9, 10), and a more ill-defined syndrome with some features akin to Chronic Fatigue Syndrome/Myalgic Encephalomyelitis (11). This latter illness, commonly characterised by intense fatigue and cognitive dysfunction (‘brain fog’) has been variably reported following other viral infections (12).

Female gender and hospitalisation during initial infection were both independently associated with an increased risk of Long COVID in our cohort. Recovery from illness causing severe pneumonia and/or intensive care stay is often prolonged and well documented to last many months (13, 14). The relationship with female gender is less well explained although confirmed in other cohorts including a large app-based study of > 4,000 individuals (1). Whether this relates to a higher risk of viral-induced immune dysregulation and auto-immunity, differences in health care utilisation, or some other mechanism is unclear. Further research is required to understand physiologic correlates of functional recovery and the role of rehabilitation intervention to assist patients with exercise capacity and dyspnoea (15).

A key strength of our study is the longitudinal assessment of symptoms, HRQOL and functional recovery performed across the spectrum of study participants at 4- and 8-months. The high rate of ongoing symptoms highlights the long-lasting and persistent nature of post-viral quality of life impact and somatic symptoms that may occur after COVID-19. Our study has several limitations. Our definition for Long COVID is conservative and based on the presence of one or more of three major symptoms of fatigue, shortness of breath or chest tightness. It does not include the full spectrum of symptoms after initial infection and is thus a probable underestimate of the true burden of ill health in this population. A third of patients were referred into the study, the reasons for which varied. In many cases this was due to concern over ongoing symptoms. To address this potential selection bias, we excluded the externally referred patients from the description of symptom prevalence and included them only for outcome analysis where we had longitudinal follow-up to compare within-individual change.

In summary, a considerable proportion of patients experience persistent symptoms after SARS-CoV-2 infection and a fifth of patient met our definition for Long COVID at 8 months. Persistent symptoms impact HRQoL and there appears to be little change between 4- and 8-months. A significant proportion of patients experience abnormal functional recovery at 8-months. The long-term significance of these findings is unknown.

## Data Availability

There is no external datasets or supplementary material online that pertain to this manuscript.

## Acknowledgements

The authors thank the research staff at the St Vincent’s Institute for Applied Medical Research.We appreciate grant support from the St Vincent’s Clinic Foundation and the Curran Foundation.

---

**Supplementary Figure 1.**
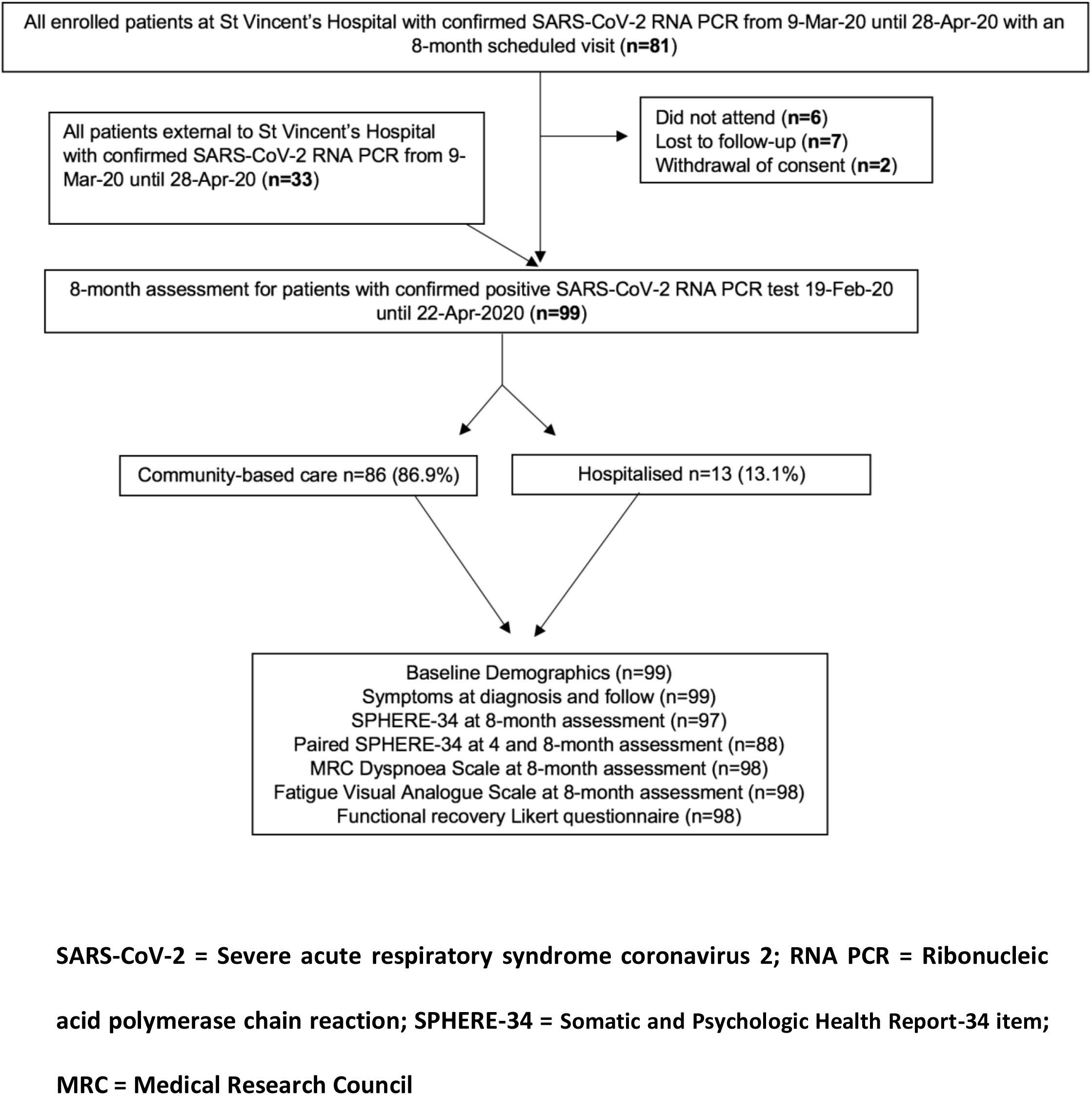
**Analytic flow diagram**

## Supplementary File

**Somatic and Psychologic Health Report-34 item (SPHERE-34). Highlighted are the somatic (SOMA) and psychologic (PSYCH) sub-scales**.

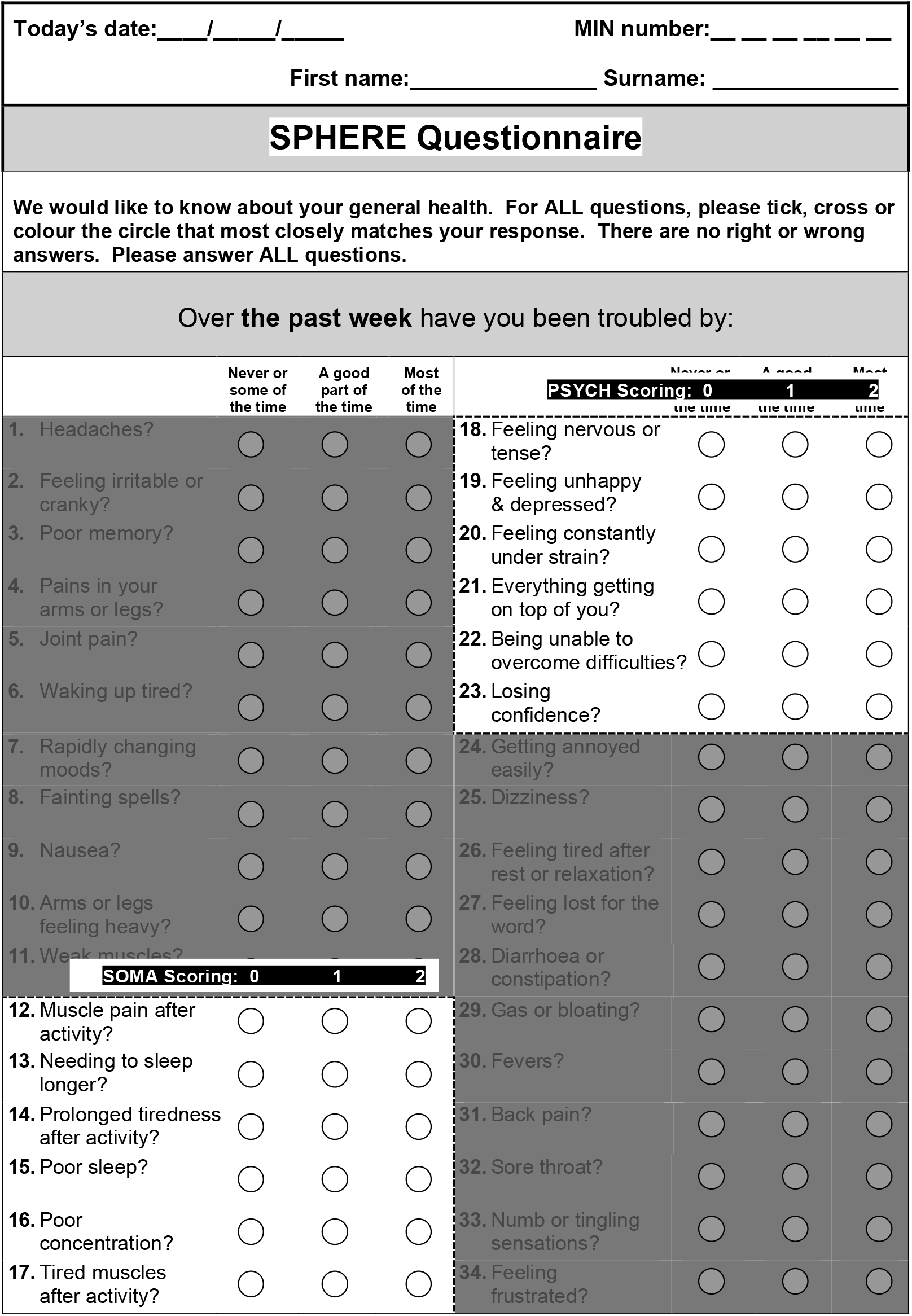

